# Retrospective Review of Blood Culture-Confirmed Cases of Enteric Fever in Navi Mumbai, India: 2014–2018

**DOI:** 10.1101/2023.05.27.23290637

**Authors:** Niniya Jayaprasad, Priyanka Borhade, Christopher LeBoa, Kashmira Date, Shrikrishna Joshi, Rahul Shimpi, Jason R. Andrews, Pankaj Bhatnagar, Stephen P. Luby, Seth A. Hoffman

## Abstract

India has one of the highest estimated burdens of enteric fever globally. Prior to implementation of Typbar-TCV^®^ typhoid conjugate vaccine (TCV) in a public sector pediatric immunization campaign in Navi Mumbai, India, we conducted a retrospective review of blood culture-confirmed cases of typhoid and paratyphoid fevers to estimate the local burden of disease. This review included all blood cultures processed at a central microbiology laboratory, serving multiple hospitals, in Navi Mumbai (January 2014–May 2018) that tested positive for either *Salmonella* Typhi or *Salmonella* Paratyphi A. Of 40,670 blood cultures analyzed, 1,309 (3.2%) were positive for S. Typhi (1,201 [92%]) or S. Paratyphi A (108 [8%]). Culture positivity was highest in the last months of the dry season (April-June). Our findings indicate a substantial burden of enteric fever in Navi Mumbai and support the importance of TCV immunization campaigns and improved water, sanitation, and hygiene.

---

Enteric fever is a disease caused by *Salmonella enterica* subspecies *enterica* serotype Typhi (S. Typhi) or Paratyphi A, B, or C (S. Paratyphi A, B, or C). Enteric fever caused an estimated 14 million cases and 136,000 deaths in 2017.^1^ The most cases occurred among children aged 5-9 years-of-age; 56% were in individuals of < 15 years.^1^ India accounted for the majority of the 2017 global burden with an estimated 8 million cases (57%) and 72,000 deaths (53%).^1^ India also has one of the highest antibiotic utilization rates in the world,^2^ alongside worsening rates of antimicrobial resistant typhoid.^3^ In the setting of increasing global prevalence of antimicrobial resistant typhoid, the World Health Organization (WHO) has prioritized typhoid vaccine delivery in conjunction with water, sanitation, and hygiene (WASH) interventions.^4^

In 2017, the WHO prequalified a new typhoid conjugate vaccine (TCV), Typbar-TCV® (Bharat Biotech International Limited, India) based on S. Typhi Vi antigen.^4^ This vaccine can be used in children as young as 6-months-old, has improved immunogenicity compared to prior typhoid vaccine generations, and has an expected longer duration of protection than unconjugated Vi antigen vaccines.^4^

In 2018, the Navi Mumbai Municipal Corporation (NMMC), the local governing body of Navi Mumbai, India, introduced the first public sector, pediatric TCV mass vaccination campaign.^5^ Navi Mumbai is located outside the industrial area of Greater Mumbai, the capital of Maharashtra State, and has a high burden of pediatric typhoid along with increasing prevalence of antimicrobial resistant typhoid isolate.^5, 6^ Prior to the wide scale implementation of Typbar-TCV® in Navi Mumbai we endeavored to understand the local burden of S. Typhi and S. Paratyphi infections.

We conducted a retrospective review of blood culture-confirmed cases of *S*. Typhi and *S*. Paratyphi A between January 2014-May 2018. Data were obtained from Dr. Joshi’s Central Clinical Microbiology Laboratory (Joshi Lab), a privately run laboratory in Navi Mumbai, that serves many local healthcare facilities. We included all blood culture samples referred to Joshi Lab during the study period, which by default included evaluation for *S*. Typhi and *S*. Paratyphi A, but not for *S*. Paratyphi B or C. All samples were submitted at the discretion of the patients’ primary providers and only samples from within NMMC boundaries were evaluated.

Joshi Lab evaluated blood culture samples using an automated blood culture system (BD-BACTEC^TM^). Further subculture was performed using sheep blood agar and MacConkey agar for differentiation of lactose fermenting from non-lactose fermenting gram-negative bacteria colonies. Conventional biochemical tests using oxidase, urease, indole, citrate, and triple sugar iron (TSI) were performed to identify and confirm pathogens (*S*. Typhi: oxidase-negative, urease-negative, indole-negative, citrate-negative, TSI K/A/H_2_S-positive/gas-negative; *S*. Paratyphi: oxidase-negative, urease-negative, indole-negative, citrate-negative, TSI K/A/H_2_S-negative/gas-positive).

A deidentified spreadsheet was shared by Joshi Lab including month and year of blood culture, and whether the blood culture was positive or negative for *S*. Typhi or *S*. Paratyphi A. A subset of positive blood cultures underwent antibiotic sensitivity testing, which included additional information about the participant (age and gender), but these antibiotic sensitivity testing results were excluded due to quality control concerns. Statistical analysis was performed in R (version 4.1.0). We evaluated the monthly blood culture *S*. Typhi and *S*. Paratyphi positivity rate compared to monthly NMMC rainfall data,^7^ and assessed sample correlations at lagged times (ccf, R version 4.1.0) to evaluate seasonal variation in the blood culture positivity rate based on rainfall amount. We used our positive cases and the Navi Mumbai 2011 census of ∼1.12 million persons^8^ to calculate crude typhoid and paratyphoid incidence rates (where complete yearly data was available from Joshi Lab, 2014-2017). Given the low sensitivity of blood culture (59%),^9^ we calculated adjusted incidences to account for missed cases [(crude incidence)*1/(1-0.59)].

Local Institutional Review Board approval was not solicited, as the study involved secondary data analysis, while the protocol for this study was approved by the Stanford University IRB (eprotocol #39627). All data provided to investigators were deidentified.

A total of 40,670 blood cultures were tested for *S*. Typhi and *S*. Paratyphi A. Of these, 1,309 (3.2%) were positive. *S*. Typhi was the predominant isolate, accounting for 1,201 blood cultures (92%), while *S*. Paratyphi A accounted for 108 blood cultures (8%).

Of 1,309 blood cultures positive for *S*. Typhi and *S*. Paratyphi A, gender information was available for 922 cases (377 females, 41%; 545 males, 59%) and of these 701 included age (<2y, 31 (4%); 2-5y, 133 (19%); >5-15y, 349 (50%); >15-45y, 175 (25%); >45y, 13 (2%); Figure 1). The median enteric fever case age was 10 years for both males and females.

**Figure 1:**
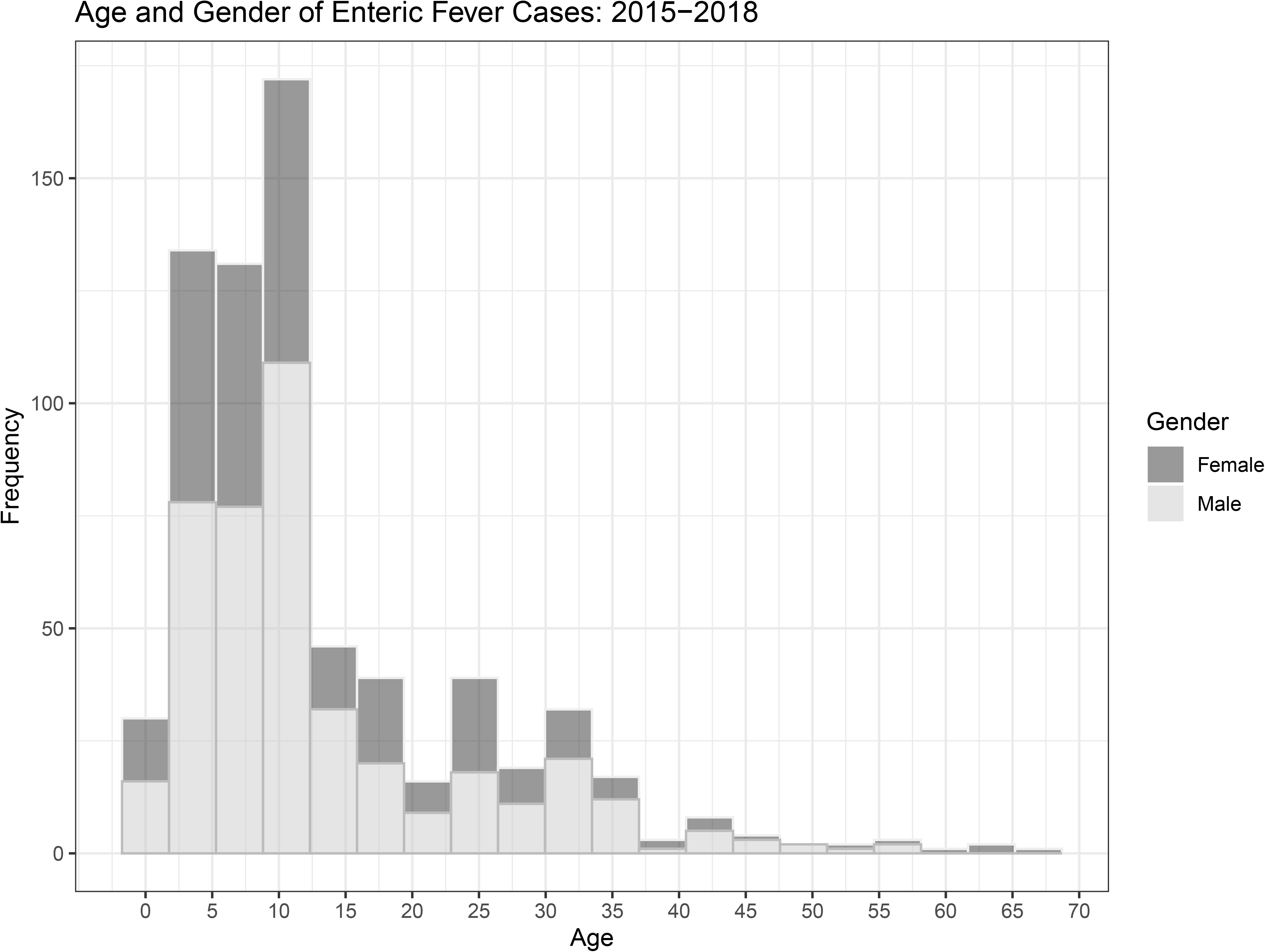
Breakdown of positive blood cultures by age and gender. Dark Grey=female, light grey=male.

The proportion of blood cultures that grew either *S*. Typhi or *S*. Paratyphi A varied by year: 2014, 3.02%; 2015, 2.57%; 2016, 3.76%; 2017, 3.19%; 2018, 4.16%. The peak proportion of blood cultures that grew either *S*. Typhi or *S*. Paratyphi A preceded peak rainfall over time by ∼2 months, suggesting an association between blood culture positivity and the terminal period of the dry season (Figure 2a; dominant cross correlation lag *h*=+2, autocorrelation=0.46, p<0.05). The percent and raw number of specimens that grew either S. Typhi or S. Paratyphi A varied by month (Figure 2a&b).

**Figure 2:**
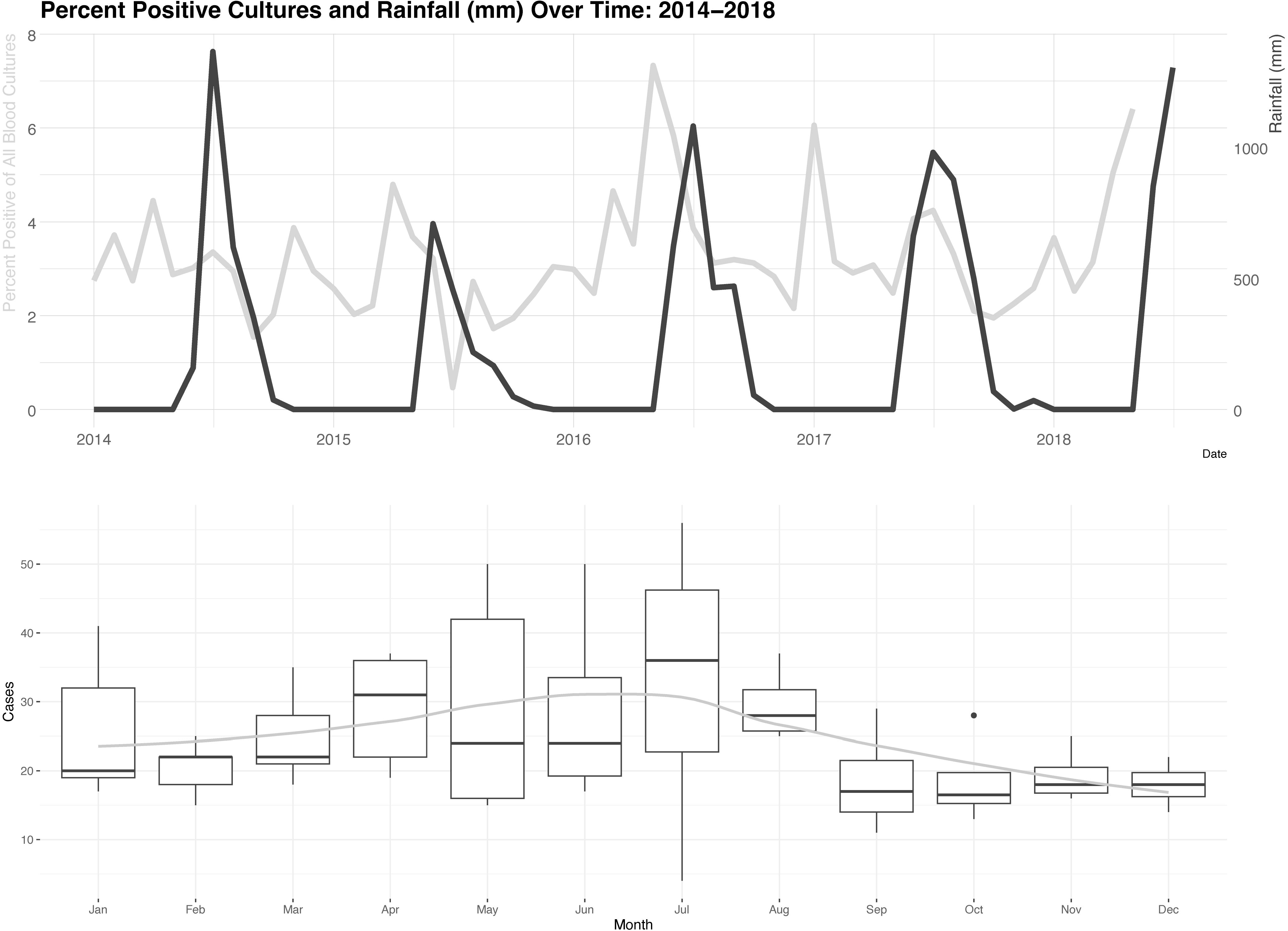
a) Percent positive blood cultures and rainfall (mm) over time: 2014-2018. Blood culture positivity in light grey, rainfall in dark grey. b) Case counts by month: 2014-2018. Smoothed conditional means in grey line.

The overall average *S*. Typhi and *S*. Paratyphi positivity rate for blood cultures performed at Joshi Lab between 2014-2018 was 3.1%, which is similar to the rate reported from Vellore, India (3.8%) over a comparable time period.^10^ A retrospective review from the Surveillance for Enteric Fever in Asia Project portrayed a much lower enteric fever blood culture positivity rate (0.53%) from 5 hospitals across India.^11^ The enteric fever blood culture positivity rate at Joshi Lab trended up over the course of the study period, while other South Asian data suggest decreased rates from 2000-2015 in India, which has been attributed to improved economic factors, sanitation, and hygiene.^12^

The majority of cases were in males (59%), which is in line with overall sex-disparities in enteric fever reporting in South Asia.^13^ For all study years a spike in case positivity preceded the monsoon’s peak rainfall by ∼2 months, except in 2017 when the highest case positivity was in January. In contrast to prior data in which South Asian enteric fever case burden was described as highest during periods of peak rainfall or just after,^14^ our sample suggests an association with the dry season in Navi Mumbai. Similarly, in a community-based case control study in Jakarta, Indonesia, there was a peak in enteric fever cases during the dry season compared to the wet season (ratio of 7:3).^15^ Depending on the source of water, during periods of water scarcity, such as the dry season, persons may be more prone to using contaminated water sources and/or the impact of a contaminant may be amplified, as has been described in cholera seasonality in South Asia.^16^

The calculated crude typhoid incidence was 23/100,000 and paratyphoid incidence was 2/100,000. The adjusted typhoid incidence was 56/100,000 and paratyphoid incidence was 5/100,000. These are likely underestimates for Navi Mumbai given blood culture results are dependent on health-care-seeking behavior, and South Asian data portray only a minority of individuals in endemic settings receive blood cultures when febrile.^17^ Even as minimum estimates from just one of Navi Mumbai’s laboratories, a typhoid incidence of 56/100,000 would be considered medium typhoid incidence.^18^ Our calculated crude incidence values are similar to recent South Asian data, in which adjusted incidences were calculated utilizing a hybrid surveillance scheme and projected 7-10x greater burden than crude estimates.^17^ If applied to our crude estimates, this would suggest an adjusted S. Typhi incidence of 161/100,000 [(23/100,000)*7] – high typhoid incidence.^18^

We utilized culture positivity over time, as prior work suggests this as a potential method of making inferences on typhoid fever incidence,^19^ though it can be confounded by other infectious diseases circulating in the community and may result in enteric fever burden underestimates. Our age and gender data are limited, as only a fraction of our total data set (age data: 922/1309, 70%; age and gender data: 701/1309, 54%) included this information, and thus may not accurately represent Navi Mumbai. Finally, we estimated crude incidence, but used >10-year-old census data, which would overestimate the crude incidence given presumed population growth since 2011. That said, blood cultures were collected at the primary provider’s discretion, which may result in missed non-severe or atypical cases and this underdiagnosis is compounded by the low sensitivity of optimally collected blood cultures (59%), made even lower with antibiotic use.^9^

This retrospective review of the results of blood cultures from just one of Navi Mumbai’s diagnostic laboratories is comparable to more complete data from throughout South Asia^10, 17^ and indicates a medium typhoid burden in Navi Mumbai at a minimum. Our data support further work to investigate mechanisms by which Navi Mumbai’s enteric fever case positivity rate is highest during the dry season. These data support the importance NMMC has placed on TCV immunization campaigns and highlight the continued importance of enteric fever surveillance and control, especially in light of increasing prevalence of antimicrobial resistant typhoid fever in South Asia and globally.

## Data Availability

All data produced in the present study are available upon reasonable request to the authors

## Acknowledgements

We thank the following organizations and individuals for their contributions to this evaluation: the Navi Mumbai Municipal Corporation leadership and staff, the Government of India Ministry of Health and Family Welfare Universal Immunization Program, State of Maharashtra Department of Public Health and Family Welfare, Indian Academy of Pediatrics Navi Mumbai Chapter, Bharat Biotech International Limited, the Indian Council of Medical Research, WHO-India National Public Health Surveillance Project, Dr. Joshi’s Central Clinical Microbiology Laboratory, Vashi (Riecha Kamboj), and the Centers for Disease Control and Prevention, Atlanta, Georgia (Dr. Kathleen Wannemuehler, Benjamin Nygren, Matt Mikoleit, and Chenhua Zhang).

## DISCLAIMER

The findings and conclusions in this report are those of the authors and do not necessarily represent the official position, policies, or views of the United States Centers for Disease Control and Prevention, or the World Health Organization.

## References

1. GBD 2017 Typhoid and Paratyphoid Collaborators, 2019. The global burden of typhoid and paratyphoid fevers: A systematic analysis for the global burden of disease study 2017. Lancet Infect Dis 19: 369–381.

2. Farooqui HH, Selvaraj S, Mehta A, Heymann DL, 2018. Community level antibiotic utilization in india and its comparison vis-à-vis european countries: Evidence from pharmaceutical sales data. PLoS One 13: e0204805.

3. Das S, Samajpati S, Ray U, Roy I, Dutta S, 2017. Antimicrobial resistance and molecular subtypes of Salmonella enterica serovar Typhi isolates from kolkata, india over a 15 years period 1998-2012. Int J Med Microbiol 307: 28–36.

4. WHO, 2019. Typhoid vaccines: Who position paper, march 2018 - recommendations. Vaccine 37: 214–216.

5. Date K, et al., 2020. Decision making and implementation of the first public sector introduction of typhoid conjugate vaccine—navi mumbai, india, 2018. Clin Infect Dis 71: S172–S178.

6. Gavhane J, Yewale V, Weekey P, Dhanya Warrior D, 2010. Enteric fever in children from navi mumbai — clinical profile, hematological features, sensitivity patterns and response to antimicrobials. Pediatr Infect Dis 2: 5–9.

7. Navi Mumbai Municipal Corporation, Rainfall data. Available at: https://www.nmmc.gov.in/navimumbai/rainfall-data. Accessed July 01, 2022.

8. Census Organization of India, Navi mumbai city census 2011 data. Available at: https://www.census2011.co.in/census/city/368-navi-mumbai.html. Accessed September 6, 2022.

9. Antillon M, Saad NJ, Baker S, Pollard AJ, Pitzer VE, 2018. The relationship between blood sample volume and diagnostic sensitivity of blood culture for typhoid and paratyphoid fever: A systematic review and meta-analysis. J Infect Dis 218: S255–s267.

10. Srinivasan M, et al., 2021. Factors predicting blood culture positivity in children with enteric fever. J Infect Dis 224: S484–s493.

11. Sur D, Barkume C, Mukhopadhyay B, Date K, Ganguly NK, Garrett D, 2018. A retrospective review of hospital-based data on enteric fever in india, 2014-2015. J Infect Dis 218: S206–s213.

12. Balaji V, Kapil A, Shastri J, Pragasam AK, Gole G, Choudhari S, Kang G, John J, 2018. Longitudinal typhoid fever trends in india from 2000 to 2015. Am J Trop Med Hyg 99: 34–40.

13. Barkume C, et al., 2018. Phase i of the surveillance for enteric fever in Asia project (seap): An overview and lessons learned. J Infect Dis 218: S188–s194.

14. Saad NJ, Lynch VD, Antillón M, Yang C, Crump JA, Pitzer VE, 2018. Seasonal dynamics of typhoid and paratyphoid fever. Sci Rep 8: 6870.

15. Vollaard AM, Ali S, van Asten HA, Widjaja S, Visser LG, Surjadi C, van Dissel JT, 2004. Risk factors for typhoid and paratyphoid fever in jakarta, indonesia. Jama 291: 2607–15.

16. Baracchini T, King AA, Bouma MJ, Rodó X, Bertuzzo E, Pascual M, 2017. Seasonality in cholera dynamics: A rainfall-driven model explains the wide range of patterns in endemic areas. Advances in Water Resources 108: 357–366.

17. Garrett DO, et al., 2022. Incidence of typhoid and paratyphoid fever in Bangladesh, Nepal, and Pakistan: Results of the surveillance for enteric fever in Asia project. Lancet Glob Health 10: e978–e988.

18. Crump JA, Luby SP, Mintz ED, 2004. The global burden of typhoid fever. Bull World Health Organ 82: 346–53.

19. Marchello CS, Dale AP, Pisharody S, Crump JA, 2019. Using hospital-based studies of community-onset bloodstream infections to make inferences about typhoid fever incidence. Trop Med Int Health 24: 1369–1383.

